# REACT-1 round 15 interim report: High and rising prevalence of SARS-CoV-2 infection in England from end of September 2021 followed by a fall in late October 2021

**DOI:** 10.1101/2021.11.03.21265877

**Authors:** Marc Chadeau-Hyam, Oliver Eales, Barbara Bodinier, Haowei Wang, David Haw, Matthew Whitaker, Caroline E. Walters, Christina Atchison, Peter J. Diggle, Andrew J. Page, Deborah Ashby, Wendy Barclay, Graham Taylor, Graham Cooke, Helen Ward, Ara Darzi, Christl A. Donnelly, Paul Elliott

## Abstract

**Background:** The third wave of COVID-19 in England coincided with the rapid spread of the Delta variant of SARS-CoV-2 from the end of May 2021. Case incidence data from the national testing programme (Pillar 2) in England may be affected by changes in testing behaviour and other biases. Community surveys may provide important contextual information to inform policy and the public health response.

**Methods:** We estimated patterns of community prevalence of SARS-CoV-2 infection in England using RT-PCR swab-positivity, demographic and other risk factor data from round 15 (interim) of the REal-time Assessment of Community Transmission-1 (REACT-1) study (round 15a, carried out from 19 to 29 October 2021). We compared these findings with those from round 14 (9 to 27 September 2021).

**Results:** During mid- to late-October 2021 (round 15a) weighted prevalence was 1.72% (1.61%, 1.84%) compared to 0.83% (0.76%, 0.89%) in September 2021 (round 14). The overall reproduction number (R) from round 14 to round 15a was 1.12 (1.11, 1.14) with increases in prevalence over this period (September to October) across age groups and regions except Yorkshire and The Humber. However, within round 15a (mid- to late-October) there was evidence of a fall in prevalence with R of 0.76 (0.65, 0.88). The highest weighted prevalence was observed among children aged 5 to 12 years at 5.85% (5.10%, 6.70%) and 13 to 17 years at 5.75% (5.02%, 6.57%). At regional level, there was an almost four-fold increase in weighted prevalence in South West from round 14 at 0.59% (0.43%,0.80%) to round 15a at 2.18% (1.84%, 2.58%), with highest smoothed prevalence at subregional level also found in South West in round 15a. Age, sex, key worker status, and presence of children in the home jointly contributed to the risk of swab-positivity. Among the 126 sequenced positive swabs obtained up until 23 October, all were Delta variant; 13 (10.3%) were identified as the AY.4.2 sub-lineage.

**Discussion:** We observed the highest overall prevalence of swab-positivity seen in the REACT-1 study in England to date in round 15a (October 2021), with a two-fold rise in swab-positivity from round 14 (September 2021). Despite evidence of a fall in prevalence from mid- to late-October 2021, prevalence remains high, particularly in school-aged children, with evidence also of higher prevalence in households with one or more children. Thus, vaccination of children aged 12 and over remains a high priority (with possible extension to children aged 5-12) to help reduce within-household transmission and disruptions to education, as well as among adults, to lessen the risk of serious disease among those infected.

## Introduction

The REal-time Assessment of Community Transmission-1 (REACT-1) study[1,2] has been tracking the spread of the COVID-19 epidemic in England approximately monthly since May 2020. The third wave of the epidemic in England took off at the end of May 2021 with the rapid rise of Delta variant and the almost complete replacement of Alpha,[3] despite the high rates of vaccination especially among people over the age of 50 years. Overall weighted prevalence of SARS-CoV-2 swab-positivity increased over four-fold between REACT-1 round 12 (20 May to 7 June 2021) and round 13 (24 June to 12 July 2021) from 0.15% to 0.63% respectively. In round 13, prevalence was highest in those aged 13 to 17 years and 18 to 24 years, while in round 14 (9 to 27 September 2021) prevalence was highest in school-aged children aged 5 to 12 years and 13 to 17 years. A key difference between these rounds is that school-aged children returned to schools in England in September 2021 with few COVID-19-related precautions in place.

Here we describe patterns of SARS-CoV-2 prevalence during October 2021 in England using interim findings from round 15 of REACT-1 (round 15a). We analysed RT-PCR swab-positivity, demographic and other risk factor data obtained from a random sample of the population of England from 19 to 29 October 2021 and compared our findings with those from September 2021 (round 14). Our aim was to obtain an up-to-date view of the progression of the epidemic in England against a backdrop of continued rollout of the vaccination programme, including (since September 2021) in children aged 12 and over, and implementation of a booster programme among adults aged 50 years and over (and some other groups). This round is being carried out during autumn associated with increased social mixing indoors as the weather becomes colder and follows the relaxation of mandatory social distancing and mask-wearing measures in England.

## Methods

### Study Population

The REACT-1 study methods are published elsewhere [2]. Briefly, prior to the current round (19 to 29 October 2021, round 15a), we completed 14 rounds of data collection over two to three weeks every month since May 2020 (except for December 2020 and August 2021). Each round, we invite a random cross-sectional sample of the population of England at ages 5 years and over based on the National Health Service (NHS) list of patients held by NHS Digital. The data sampling method was changed during the study. Prior to round 12 (May to June 2021) we aimed to achieve approximately equal numbers of participants for the n=315 lower-tier local authorities (LTLAs) in England (Isles of Scilly combined with Cornwall and the City of London with Westminster). From round 12 onwards, this approach was modified to invite a random sample of the population in proportion to population-size at LTLA level, thus increasing the numbers sampled in urban areas with higher population density and reducing the numbers in sparsely populated rural areas. This change, however, should not have affected prevalence estimates as these are weighted to be representative of England as a whole. We also changed the method of collection of swabs (for RT-PCR testing) from round 14 (September 2021), from dry swabs in prior rounds to ‘wet’ swabs (in saline), and to use of the priority postal service instead of courier pick-up (on cold chain) in previous rounds. In round 14, we tested wet swabs pick-up by courier without cold chain versus use of the priority postal service and did not detect a difference between the two methods [4].

### RT-PCR testing, demographic data and questionnaire

We asked participants to provide a self-administered throat and nose swab (or their parent/guardian to administer the swab for children at ages 12 years and under) using written and video instructions. Swabs were sent for RT-PCR to a single laboratory, with a positive test for SARS-CoV-2 infection recorded either if two gene targets (N gene and E gene) were detected or if N gene was detected with cycle threshold (Ct) value below 37.

From the NHS register, we obtained information on age, sex and residential location with questionnaire data providing additional information on demography derived from an online or telephone questionnaire [5].

### Viral genome sequencing

Samples testing positive (N gene Ct values < 34 and sufficient volume) were sent for viral genome sequencing at the Quadram Institute, Norwich, UK, using the ARTIC protocol [6] for viral RNA amplification and CoronaHiT for preparation of sequencing libraries [7]. Analysis of sequencing data used the ARTIC bioinformatic pipeline [8] and we assigned lineages using PangoLEARN[9].

### Statistical Analyses

We carried out the statistical analyses in R [10]. We calculated unweighted prevalence of swab-positivity (from RT-PCR) and weighted prevalence using rim weighting [11] to provide estimates that were representative of the population of England as a whole.

We analysed trends in swab-positivity over time using an exponential model of growth or decay, with the assumption that numbers of positive samples from the total number of samples per day arose from a binomial distribution. We used day of swabbing where available or day of first scan of the sample by the Post Office otherwise. We used a bivariate No-U-Turn Sampler to estimate posterior credible intervals assuming uniform prior distributions on the probability of swab-positivity on day of swabbing and the growth rate [12]. We estimated the reproduction number R for all participants and at all ages, and for specific sub-groups, assuming a gamma-distributed generation time with shape parameter, n=2.29 and rate parameter *β*=0.36 (corresponding to a mean generation time of 6.29 days) [13].

To visualise trends over time, we fit a Bayesian penalised-spline (P-spline) model [14] to the daily data using a No-U-Turn Sampler in logit space. We segmented the data into approximately 5-day sections by regularly spaced knots, with further knots included beyond the study period to minimise edge effects. We also fit P-splines separately to three broad age groups (17 years and under, 18 to 54 years, 55 years and over) with a smoothing parameter obtained from the model fit to all data.

The daily growth rate of the proportion of AY.4.2 sub-lineage relative to all other lineages was estimated by fitting a Bayesian logistic regression model assuming a Binomial likelihood in the daily proportion of AY.4.2.

We estimated smoothed LTLA prevalence from a subsample of N=15 individuals per LTLA by taking the median number of nearest neighbours (i.e. within a 30km radius) of each individual, calculating neighbourhood prevalence for that individual, then taking the mean neighbourhood prevalence across all N=15 individuals per LTLA. We defined individual neighbourhood prevalence as the number of nearest neighbours with positive tests divided by the total number of nearest neighbours.

## Results

### Descriptive statistics and prevalence estimates

A total of 859,184 participants were invited to participate in round 15. Amongst invited participants 143,193 (16.7%) registered of whom 67,208 (46.9%) provided a swab with a valid result from RT-PCR up to 29 October 2021 (round 15a) (Supplementary Table 1). Of these 67,208 valid swabs in round 15a, 1,021 were positive giving a weighted prevalence of 1.72% (1.61%, 1.84%). The weighted prevalence in round 15a is more than two-fold higher than that estimated in round 14 at 0.83% (0.76%, 0.89%).

The highest weighted prevalence by age in round 15a was observed in those aged 5-12 years at 5.85% (5.10%, 6.70%) and those aged 13-17 years at 5.75% (5.02%, 6.57%) (Table 1a, Figure 1-A). The next highest weighted prevalences by age were found in those aged 45-54 years at 1.53% (1.29%, 1.80%) and 35-44 years at 1.48% (1.21%, 1.80%). Weighted prevalence was 0.82% (0.68%, 0.99%) in those aged 65-74 years and 0.67% (0.50%, 0.89%) in those 75 years and over, both representing increases of approximately two-fold from round 14.

**Table 1a.**
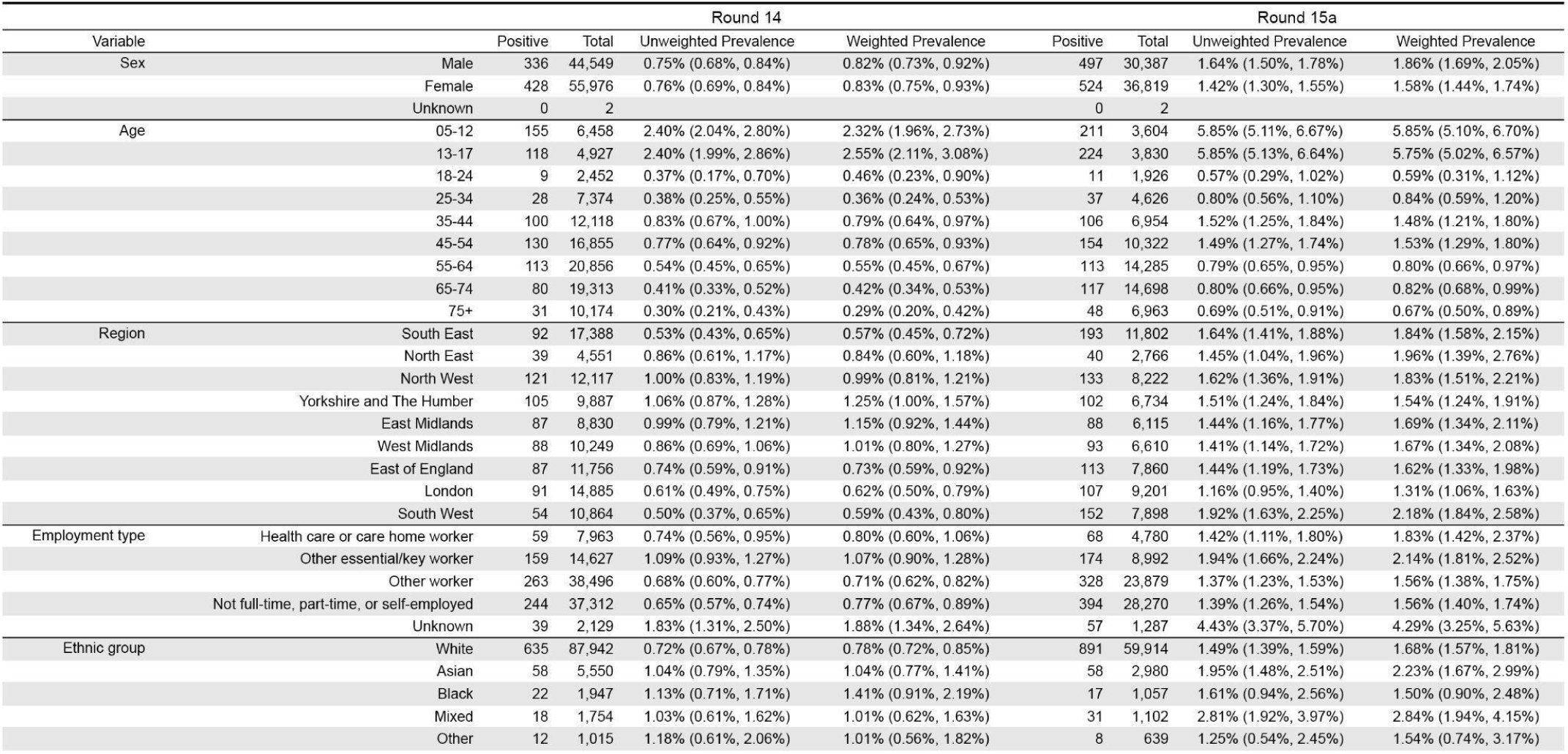
Unweighted and weighted prevalence of swab-positivity in round 14 and round 15a by sex, age, region, employment type, and ethnic group.

**Figure 1.**
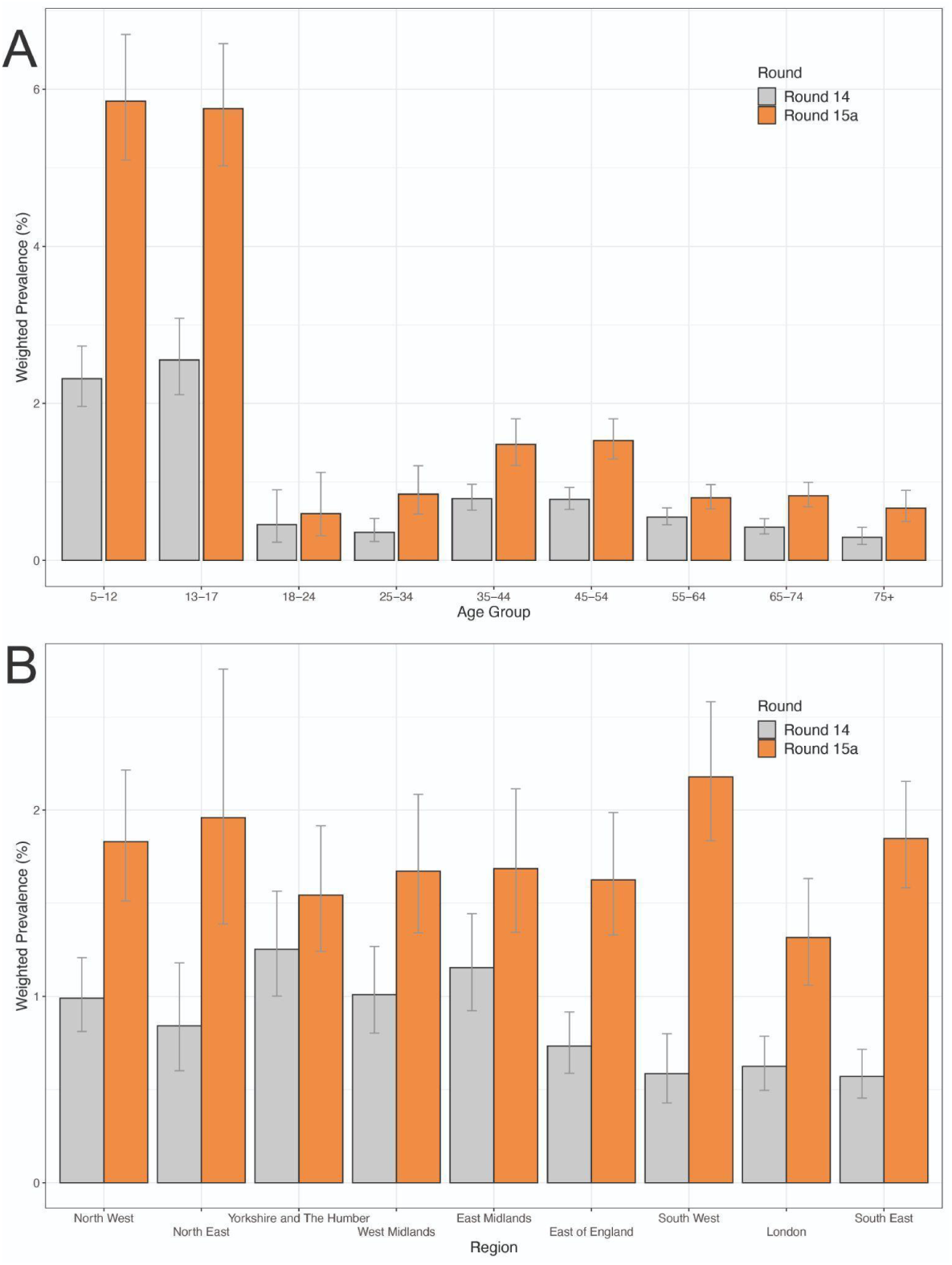
Weighted prevalence of swab-positivity in round 14 (grey) and round 15a (orange) by (A) age group and (B) region. Bars show the prevalence point estimates, and the vertical lines represent the 95% confidence intervals.

The highest weighted prevalence in round 15a by region was found in South West at 2.18% (1.84%, 2.58%) increasing almost four-fold from round 14 at 0.59% (0.43%, 0.80%) (Table 1a, Figure 1-B). Across rounds 14 and 15a, the weighted prevalence was found to be growing with >0.99 posterior probability that R>1 in all regions except Yorkshire and The Humber (Table 2). However, within round 15a, there was a fall in prevalence in East Midlands with R=0.51 (0.26, 0.86), East of England with R=0.61 (0.35, 0.97), and South West with R=0.59 (0.36, 0.90).

At LTLA level, we observed a general increase in the smoothed estimates of prevalence between round 14 and round 15a (Figure 2). The ten LTLAs with the highest smoothed prevalence nationally (≥2.36%) in round 15a were all in South West: Cheltenham, Stroud, Swindon, Gloucester, Cotswold, South Gloucestershire, Wiltshire, Tewkesbury, Bath and North East Somerset, and City of Bristol.

**Figure 2.**
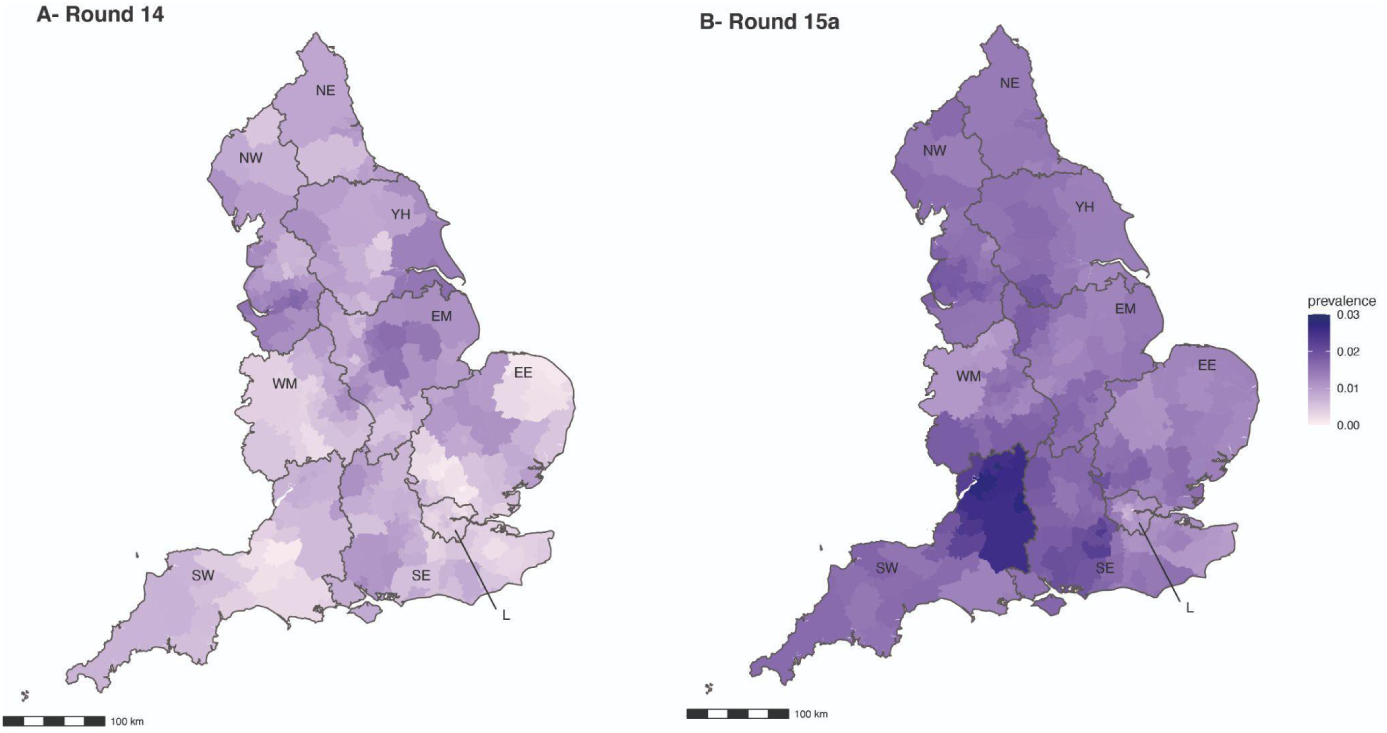
Neighbourhood smoothed average prevalence by lower tier local authority area for (A) round 14 and (B) round 15a. Neighbourhood prevalence calculated from nearest neighbours (the median number of neighbours within 30 km in the study). Average neighbourhood prevalence displayed for individual lower-tier local authorities. Regions: NE = North East, NW = North West, YH = Yorkshire and The Humber, EM = East Midlands, WM = West Midlands, EE = East of England, L = London, SE = South East, SW = South West

In round 15a, we observed the highest weighted prevalence in: those reporting any of the following common COVID-19 symptoms in the month prior to testing (loss or change of sense of smell or taste, fever, new persistent cough) at 8.69% (7.90%, 9.55%) compared to 0.71% (0.62%, 0.81%) in those without symptoms; those in contact with a confirmed COVID-19 case at 10.1% (9.19%, 11.2%) compared to 0.83% (0.74%, 0.92%) for people without such contact; larger households including 5 or more people at 3.68% (3.05%, 4.43%) and 6 or more people at 3.03% (2.24%, 4.09%) compared to 0.78% (0.62%, 0.98%) in single-person households; and households with one or more children at 3.09% (2.80%, 3.41%) compared to 0.75% (0.67%, 0.84%) in households without children (Table 1b).

**Table 1b.** Unweighted and weighted prevalence of swab-positivity in round 14 and round 15a by household size, contact with a COVID-19 case, symptom status and neighbourhood deprivation.

|  |  | Round 14 |  |  |  | Round 15a |  |  |  |
| --- | --- | --- | --- | --- | --- | --- | --- | --- | --- |
| Variable |  | Positive | Total | Unweighted Prevalence | Weighted Prevalence | Positive | Total | Unweighted Prevalence | Weighted Prevalence |
| Household size | 1 | 57 | 16,613 | 0.34% (0.26%, 0.44%) | 0.33% (0.25%, 0.44%) | 86 | 11,583 | 0.74% (0.59%, 0.92%) | 0.78% (0.62%, 0.98%) |
|  | 2 | 174 | 39,044 | 0.45% (0.38%, 0.52%) | 0.46% (0.39%, 0.54%) | 242 | 28,122 | 0.86% (0.76%, 0.98%) | 0.89% (0.77%, 1.02%) |
|  | 3 | 150 | 17,235 | 0.87% (0.74%, 1.02%) | 0.93% (0.78%, 1.10%) | 204 | 11,328 | 1.80% (1.56%, 2.06%) | 1.96% (1.68%, 2.28%) |
|  | 4 | 250 | 19,154 | 1.31% (1.15%, 1.48%) | 1.32% (1.15%, 1.51%) | 309 | 11,008 | 2.81% (2.51%, 3.13%) | 2.96% (2.63%, 3.34%) |
|  | 5 | 89 | 6,057 | 1.47% (1.18%, 1.81%) | 1.41% (1.12%, 1.76%) | 132 | 3,641 | 3.63% (3.04%, 4.28%) | 3.68% (3.05%, 4.43%) |
|  | 6+ | 44 | 2,424 | 1.82% (1.32%, 2.43%) | 1.75% (1.24%, 2.46%) | 48 | 1,526 | 3.15% (2.33%, 4.15%) | 3.03% (2.24%, 4.09%) |
| Number of children in the household | 0 | 276 | 66,025 | 0.42% (0.37%, 0.47%) | 0.40% (0.35%, 0.46%) | 349 | 46,236 | 0.75% (0.68%, 0.84%) | 0.75% (0.67%, 0.84%) |
|  | 1+ | 378 | 28,659 | 1.32% (1.19%, 1.46%) | 1.37% (1.22%, 1.52%) | 463 | 16,510 | 2.80% (2.56%, 3.07%) | 3.09% (2.80%, 3.41%) |
|  | Unknown | 110 | 5,843 | 1.88% (1.55%, 2.26%) | 2.06% (1.69%, 2.51%) | 209 | 4,462 | 4.68% (4.08%, 5.35%) | 4.68% (4.07%, 5.39%) |
| COVID case contact | No | 325 | 80,587 | 0.40% (0.36%, 0.45%) | 0.43% (0.38%, 0.49%) | 406 | 54,669 | 0.74% (0.67%, 0.82%) | 0.83% (0.74%, 0.92%) |
|  | Yes, contact with a confirmed/tested COVID-19 case | 293 | 4,044 | 7.25% (6.47%, 8.09%) | 7.35% (6.50%, 8.31%) | 437 | 4,597 | 9.51% (8.67%, 10.39%) | 10.14% (9.19%, 11.18%) |
|  | Yes, contact with a suspected COVID-19 case | 38 | 1,058 | 3.59% (2.55%, 4.90%) | 3.80% (2.70%, 5.32%) | 50 | 1,015 | 4.93% (3.68%, 6.44%) | 5.25% (3.91%, 7.02%) |
|  | Unknown | 108 | 14,838 | 0.73% (0.60%, 0.88%) | 0.79% (0.65%, 0.97%) | 128 | 6,927 | 1.85% (1.54%, 2.19%) | 2.01% (1.66%, 2.43%) |
| Symptom status | Classic COVID symptoms* | 359 | 4,963 | 7.23% (6.53%, 7.99%) | 6.85% (6.12%, 7.68%) | 486 | 5,601 | 8.68% (7.95%, 9.44%) | 8.69% (7.90%, 9.55%) |
|  | Other symptoms | 108 | 11,337 | 0.95% (0.78%, 1.15%) | 0.95% (0.77%, 1.17%) | 148 | 10,532 | 1.41% (1.19%, 1.65%) | 1.56% (1.31%, 1.86%) |
|  | No symptoms | 190 | 69,446 | 0.27% (0.24%, 0.32%) | 0.31% (0.27%, 0.37%) | 262 | 44,167 | 0.59% (0.52%, 0.67%) | 0.71% (0.62%, 0.81%) |
|  | Unknown | 107 | 14,781 | 0.72% (0.59%, 0.87%) | 0.79% (0.64%, 0.97%) | 125 | 6,908 | 1.81% (1.51%, 2.15%) | 1.97% (1.62%, 2.38%) |
| Deprivation | 1 Most deprived | 119 | 11,777 | 1.01% (0.84%, 1.21%) | 0.98% (0.81%, 1.20%) | 114 | 7,560 | 1.51% (1.25%, 1.81%) | 1.69% (1.39%, 2.05%) |
|  | 2 | 118 | 16,962 | 0.70% (0.58%, 0.83%) | 0.75% (0.61%, 0.92%) | 162 | 10,949 | 1.48% (1.26%, 1.72%) | 1.72% (1.45%, 2.03%) |
|  | 3 | 142 | 21,117 | 0.67% (0.57%, 0.79%) | 0.75% (0.63%, 0.90%) | 205 | 14,254 | 1.44% (1.25%, 1.65%) | 1.64% (1.41%, 1.90%) |
|  | 4 | 178 | 24,102 | 0.74% (0.63%, 0.85%) | 0.76% (0.65%, 0.89%) | 239 | 16,367 | 1.46% (1.28%, 1.66%) | 1.71% (1.49%, 1.96%) |
|  | 5 Least deprived | 207 | 26,569 | 0.78% (0.68%, 0.89%) | 0.90% (0.77%, 1.04%) | 301 | 18,078 | 1.67% (1.48%, 1.86%) | 1.85% (1.64%, 2.08%) |

\* Classic COVID symptoms: loss or change of sense of smell or taste, fever, new persistent cough

### Epidemic growth estimates

The P-spline regression fit to data from all rounds of REACT-1 showed an increasing weighted prevalence of swab-positivity from round 14 to round 15a followed by a fall during round 15a (Figure 3-A). The posterior distribution of the estimated epidemic curve suggested that the peak in weighted prevalence was reached on around 19 to 20 October (14 to 23 October) (Figure 3-B).

**Figure 3.**
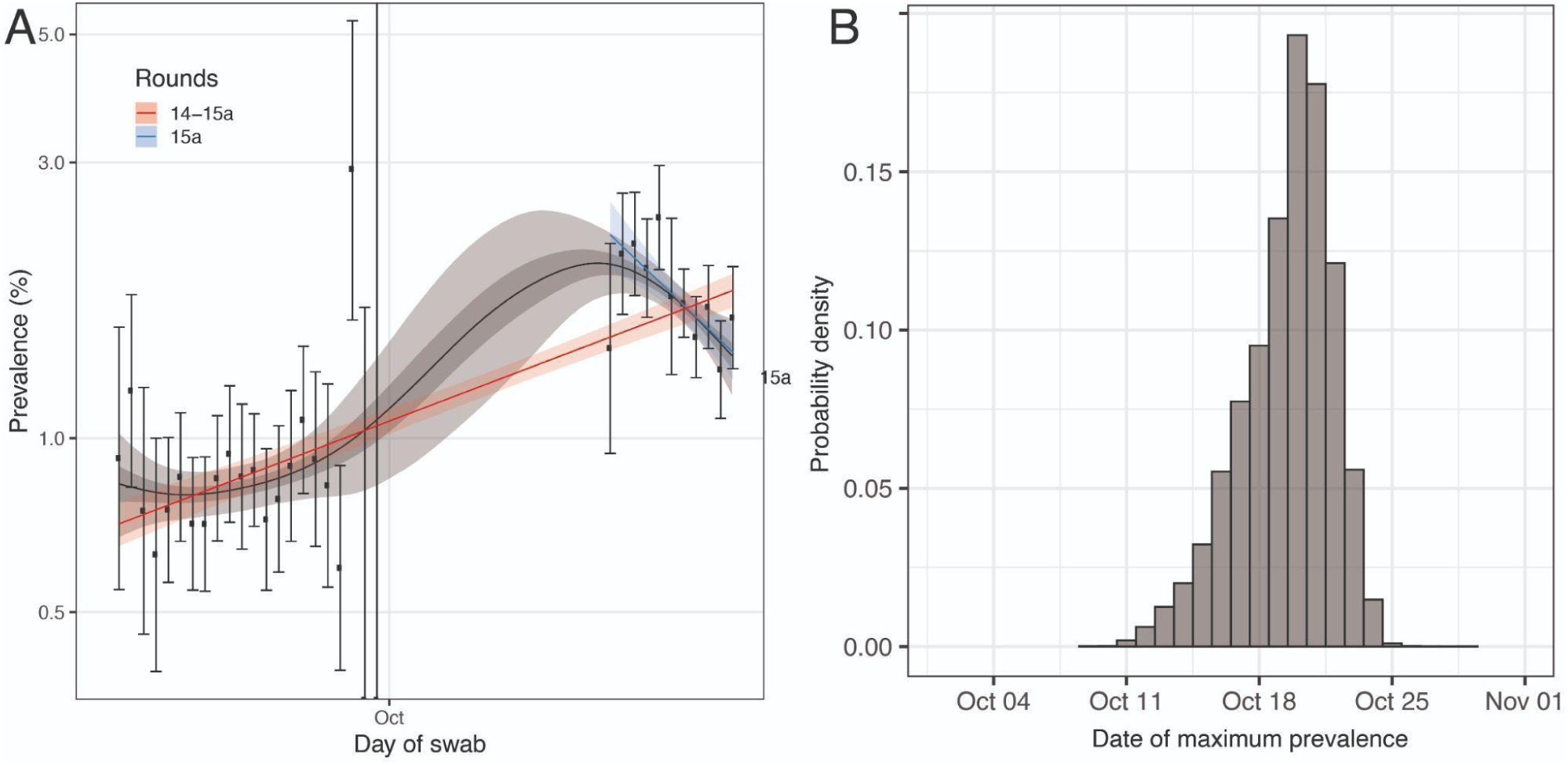
(A) Comparison of an exponential model fit to round 14-15al (red) and round 15a only (blue) and a P-spline model fit to all rounds of REACT-1 (black, shown here only for round 14 and 15a). Shaded red region shows the 95% posterior credible interval for the exponential model, and the shaded grey region shows 50% (dark grey) and 95% (light grey) posterior credible interval for the P-spline model. Results are presented for each day (X axis) of sampling for round 14 and round 15a and the prevalence of infection is shown (Y axis) on a log scale. Unweighted observations (black dots) and 95% confidence intervals (vertical lines) are also shown. (B) Probability density for the estimated date at which weighted prevalence, as estimated from the P-spline model, was at a maximum during the period of round 14 to round 15a.

Exponential models fit to data from rounds 14 and 15a estimated a reproduction number R of 1.12 (1.11, 1.14) with posterior probability that R>1 of greater than 0.99 (Table 2). Weighted prevalence increased between rounds 14 and 15a across all ages (Figure 1-A) with R ranging from 1.09 (1.06, 1.13) in those aged 55 years and over to 1.15 (1.13, 1.18) in those aged 17 years and under and posterior probability that R>1 greater than 0.99 (Table 2).

**Table 2.**
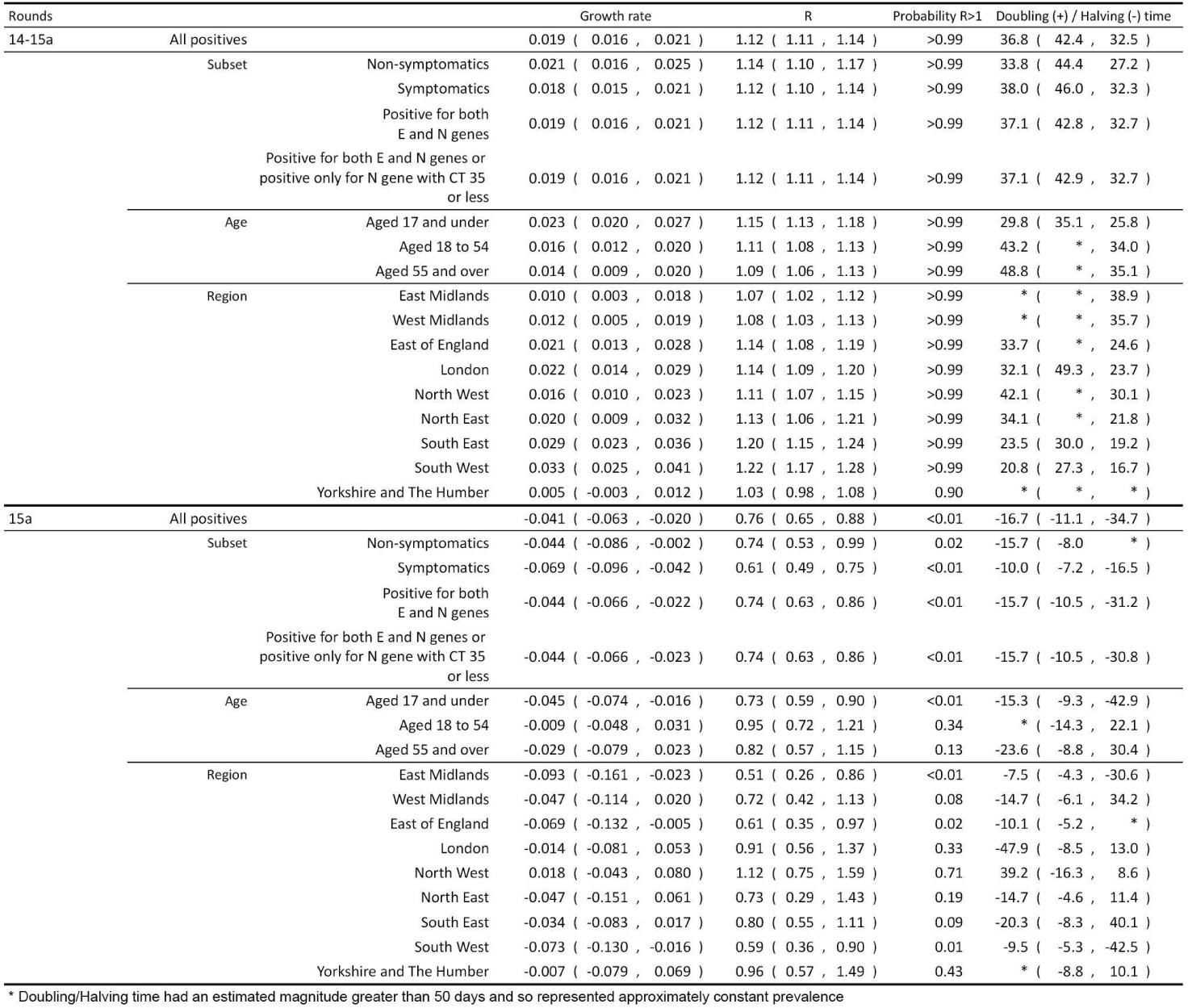
Table of growth rates, reproduction numbers and doubling/halving times from exponential model fits on data from round 14 and round 15a (top table), and round 15a only (bottom table)

When fit to data from round 15a only, the exponential model indicated a fall in prevalence during October 2021 with an R of 0.76 (0.65, 0.88) and posterior probability that R>1 less than 0.01, and this was also found among those 17 years and younger with R of 0.73 (0.59, 0.90), with suggestion of similar patterns at older ages (Table 2, Supplementary Figure 1).

### Logistic regression

Multiple logistic regression (Table 3) showed a decreased risk of swab-positivity in females with an odds ratio (OR) of 0.87 (0.77, 0.99) in round 15a when adjusting for all other covariates and an increased risk in “other essential/key workers” (i.e. not including health and care workers) compared to other workers with an OR of 1.25 (1.03, 1.51). Mutually adjusted OR for households with one or more children was 2.09 (1.63, 2.68) in round 15a compared to households without children, while the excess risk in larger compared to smaller households apparent in models adjusted for sex and age was no longer evident with adjustment for other covariates.

**Table 3.**
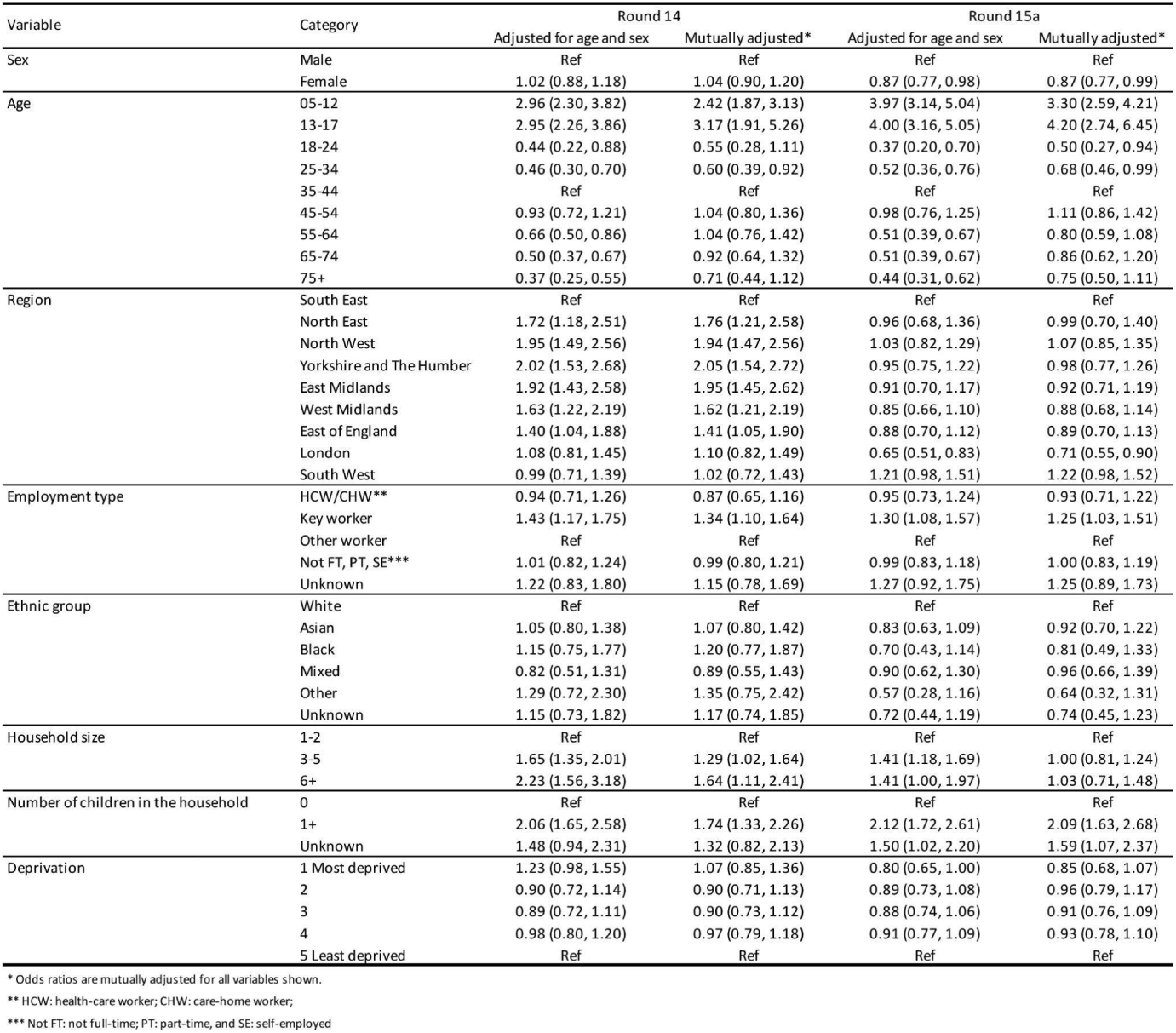
Multiple logistic regression for rounds 14 and 15a. Results are presented as estimated Odds Ratios (95% confidence interval) adjusted for age and sex and additionally for all other variables (mutually adjusted OR).

| Variable | Category | Round 14 |  | Round 15a |  |
| --- | --- | --- | --- | --- | --- |
|  |  | Adjusted for age and sex | Mutually adjusted* | Adjusted for age and sex | Mutually adjusted* |
| Sex | Male | Ref | Ref | Ref | Ref |
|  | Female | 1.02 (0.88, 1.18) | 1.04 (0.90, 1.20) | 0.87 (0.77, 0.98) | 0.87 (0.77, 0.99) |
| Age | 05-12 | 2.96 (2.30, 3.82) | 2.42 (1.87, 3.13) | 3.97 (3.14, 5.04) | 3.30 (2.59, 4.21) |
|  | 13-17 | 2.95 (2.26, 3.86) | 3.17 (1.91, 5.26) | 4.00 (3.16, 5.05) | 4.20 (2.74, 6.45) |
|  | 18-24 | 0.44 (0.22, 0.88) | 0.55 (0.28, 1.11) | 0.37 (0.20, 0.70) | 0.50 (0.27, 0.94) |
|  | 25-34 | 0.46 (0.30, 0.70) | 0.60 (0.39, 0.92) | 0.52 (0.36, 0.76) | 0.68 (0.46, 0.99) |
|  | 35-44 | Ref | Ref | Ref | Ref |
|  | 45-54 | 0.93 (0.72, 1.21) | 1.04 (0.80, 1.36) | 0.98 (0.76, 1.25) | 1.11 (0.86, 1.42) |
|  | 55-64 | 0.66 (0.50, 0.86) | 1.04 (0.76, 1.42) | 0.51 (0.39, 0.67) | 0.80 (0.59, 1.08) |
|  | 65-74 | 0.50 (0.37, 0.67) | 0.92 (0.64, 1.32) | 0.51 (0.39, 0.67) | 0.86 (0.62, 1.20) |
|  | 75+ | 0.37 (0.25, 0.55) | 0.71 (0.44, 1.12) | 0.44 (0.31, 0.62) | 0.75 (0.50, 1.11) |
| Region | South East | Ref | Ref | Ref | Ref |
|  | North East | 1.72 (1.18, 2.51) | 1.76 (1.21, 2.58) | 0.96 (0.68, 1.36) | 0.99 (0.70, 1.40) |
|  | North West | 1.95 (1.49, 2.56) | 1.94 (1.47, 2.56) | 1.03 (0.82, 1.29) | 1.07 (0.85, 1.35) |
|  | Yorkshire and The Humber | 2.02 (1.53, 2.68) | 2.05 (1.54, 2.72) | 0.95 (0.75, 1.22) | 0.98 (0.77, 1.26) |
|  | East Midlands | 1.92 (1.43, 2.58) | 1.95 (1.45, 2.62) | 0.91 (0.70, 1.17) | 0.92 (0.71, 1.19) |
|  | West Midlands | 1.63 (1.22, 2.19) | 1.62 (1.21, 2.19) | 0.85 (0.66, 1.10) | 0.88 (0.68, 1.14) |
|  | East of England | 1.40 (1.04, 1.88) | 1.41 (1.05, 1.90) | 0.88 (0.70, 1.12) | 0.89 (0.70, 1.13) |
|  | London | 1.08 (0.81, 1.45) | 1.10 (0.82, 1.49) | 0.65 (0.51, 0.83) | 0.71 (0.55, 0.90) |
|  | South West | 0.99 (0.71, 1.39) | 1.02 (0.72, 1.43) | 1.21 (0.98, 1.51) | 1.22 (0.98, 1.52) |
| Employment type | HCW/CHW** | 0.94 (0.71, 1.26) | 0.87 (0.65, 1.16) | 0.95 (0.73, 1.24) | 0.93 (0.71, 1.22) |
|  | Key worker | 1.43 (1.17, 1.75) | 1.34 (1.10, 1.64) | 1.30 (1.08, 1.57) | 1.25 (1.03, 1.51) |
|  | Other worker | Ref | Ref | Ref | Ref |
|  | Not FT, PT, SE*** | 1.01 (0.82, 1.24) | 0.99 (0.80, 1.21) | 0.99 (0.83, 1.18) | 1.00 (0.83, 1.19) |
|  | Unknown | 1.22 (0.83, 1.80) | 1.15 (0.78, 1.69) | 1.27 (0.92, 1.75) | 1.25 (0.89, 1.73) |
| Ethnic group | White | Ref | Ref | Ref | Ref |
|  | Asian | 1.05 (0.80, 1.38) | 1.07 (0.80, 1.42) | 0.83 (0.63, 1.09) | 0.92 (0.70, 1.22) |
|  | Black | 1.15 (0.75, 1.77) | 1.20 (0.77, 1.87) | 0.70 (0.43, 1.14) | 0.81 (0.49, 1.33) |
|  | Mixed | 0.82 (0.51, 1.31) | 0.89 (0.55, 1.43) | 0.90 (0.62, 1.30) | 0.96 (0.66, 1.39) |
|  | Other | 1.29 (0.72, 2.30) | 1.35 (0.75, 2.42) | 0.57 (0.28, 1.16) | 0.64 (0.32, 1.31) |
|  | Unknown | 1.15 (0.73, 1.82) | 1.17 (0.74, 1.85) | 0.72 (0.44, 1.19) | 0.74 (0.45, 1.23) |
| Household size | 1-2 | Ref | Ref | Ref | Ref |
|  | 3-5 | 1.65 (1.35, 2.01) | 1.29 (1.02, 1.64) | 1.41 (1.18, 1.69) | 1.00 (0.81, 1.24) |
|  | 6+ | 2.23 (1.56, 3.18) | 1.64 (1.11, 2.41) | 1.41 (1.00, 1.97) | 1.03 (0.71, 1.48) |
| Number of children in the household | 0 | Ref | Ref | Ref | Ref |
|  | 1+ | 2.06 (1.65, 2.58) | 1.74 (1.33, 2.26) | 2.12 (1.72, 2.61) | 2.09 (1.63, 2.68) |
|  | Unknown | 1.48 (0.94, 2.31) | 1.32 (0.82, 2.13) | 1.50 (1.02, 2.20) | 1.59 (1.07, 2.37) |
| Deprivation | 1 Most deprived | 1.23 (0.98, 1.55) | 1.07 (0.85, 1.36) | 0.80 (0.65, 1.00) | 0.85 (0.68, 1.07) |
|  | 2 | 0.90 (0.72, 1.14) | 0.90 (0.71, 1.13) | 0.89 (0.73, 1.08) | 0.96 (0.79, 1.17) |
|  | 3 | 0.89 (0.72, 1.11) | 0.90 (0.73, 1.12) | 0.88 (0.74, 1.06) | 0.91 (0.76, 1.09) |
|  | 4 | 0.98 (0.80, 1.20) | 0.97 (0.79, 1.18) | 0.91 (0.77, 1.09) | 0.93 (0.78, 1.10) |
|  | 5 Least deprived | Ref | Ref | Ref | Ref |

\* Odds ratios are mutually adjusted for all variables shown.
\*\* HCW: health-care worker; CHW: care-home worker;
\*\*\* Not FT: not full-time; PT: part-time, and SE: self-employed

### Viral sequencing

Sequencing of the positive samples collected up to 23 October 2021 has thus far yielded 126 lineages, which were all Delta or Delta sub-lineage variants (Supplementary Table 2). Of these, 60.3% (51.6%, 68.4%, N=76) were AY.4, with AY.4.2 representing 10.3% (6.13%, 16.9%, N=13) of all lineages. This represents a daily growth rate in the proportion of AY.4.2 (using Y145H mutation as a proxy in round 14) of 3.20% (1.02%, 5.38%) from round 14 onwards.

## Discussion

In this interim report from the fifteenth round of the REACT-1 study (round 15a, 19 to 29 October 2021) we observed the highest weighted prevalence of SARS-CoV-2 swab-positivity in England of any round of REACT-1 since the study commenced in May 2020. The overall prevalence of 1.72% equates to approximately 1,285,000 people in England infected with SARS-CoV-2 on any one day during mid- to late-October 2021, assuming 75% sensitivity to detect virus from a single swab. Although we observed a fall in swab-positivity towards the end of round 15a, similar to what has been seen in the routine national testing programme (Pillar 2), we have shown previously that SARS-CoV-2 prevalence is volatile. Specifically, we observed a fall and then rise in swab-positivity during the same period in 2020 (round 6, mid-October to early November 2020) [15] suggesting that we should be cautious about any extrapolation of the current downward trend going forward. This is especially so given that the fall in swab-positivity (seen across the entire age range) coincides with the autumn half-term school holidays in England when social mixing patterns will have changed.

In this regard, we found the highest prevalence to be in school-aged children, reaching nearly six percent (one in 17) in October 2021. We also reported that school-aged children had the highest prevalence in round 14 (9 to 27 September 2021). These findings likely reflect more than a month of increased social mixing of children, including indoors, as they attended school for the autumn term. The age groups with the next highest prevalences in round 15a corresponded to the parental ages (35 to 44 years and 45 to 54 years) of many school-aged children, and the presence of one or more children in the home was associated with an increased risk of swab-positivity. Larger households were no longer associated with increased risks of swab-positivity once other covariates were adjusted for, including the presence of one or more children in the home. It therefore seems likely that the high prevalence of infection among school-aged children has helped drive infection rates among other age groups including the most vulnerable (ages 65 years and over) where swab-positivity rates approximately doubled between rounds 14 and 15a (September to October 2021).

Household transmission studies have likewise indicated that household contacts are a source of infection among household members, including those who have been vaccinated [16]. The rollout of vaccination to older teenagers (aged 16 and 17 years) from August 2021 and children aged 12 to 15 years from September 2021 is likely to reduce rates of infection of, and transmission from, these secondary school-aged children - although under the current policy the 12 to 15-year old group is scheduled to receive only one dose and those under 12 years remain unvaccinated.

At regional level, we found increasing prevalence of swab-positivity from September to October 2021 in all regions except for Yorkshire and The Humber, with a nearly four-fold increase in South West over that period, resulting in prevalence of over two percent in South West in October 2021. The finding of the strongest growth from round 14 to round 15a in South West, and highest weighted prevalence in that region in round 15a, might be related to widespread reports of people who tested positive using lateral flow tests going on to test negative using RT-PCR tests. Reports specifically tied false negative RT-PCR results to a COVID-19 test laboratory in Wolverhampton which received many samples from South West [17].

As in our previous reports (round 13 [3] and round 14 [4]), we found that all sequenced swabs were Delta variant and its sub-lineages, indicating, in our own data, complete replacement of Alpha and other variants by Delta in England. Whereas in round 14 we found that 4.63% (3.07%, 6.91%) of the sequenced swabs were the AY.4.2 sub-lineage [4], in round 15a, 10.3% (6.13%, 16.9%) were AY.4.2, consistent with the UK Health Security Agency (UKHSA) report that in the week 18 October 2021 to 24 October 2021 AY.4.2 accounted for 11.3% of all sequences generated, on an increasing trajectory [18].

As with any study, the REACT-1 study has limitations. We cannot meaningfully calculate a response rate for round 15 since it is not yet complete, but between rounds 1 (May 2020) and 14 (September 2021) the response rate declined from approximately 30% to approximately 12%. Although we used rim weighting to adjust our prevalence estimates for differential response by age, sex, deprivation, LTLA counts and ethnicity, it remains possible that our estimates are not fully representative of the population as a whole. The data on participants who consented for their REACT-1 data to be linked to their NHS records which include data from the COVID-19 immunisation programme are not yet available for round 15.

Surveillance in England indicates that influenza incidence was low up to 26 October 2021 (the most recent data reported by UKHSA [19]), although it will almost certainly increase in coming weeks unless strict social distancing were to be reinstated. For viral infections, co-infections with other bacterial and viral pathogens can increase the risks of complications and mortality. The combined effects of SARS-CoV-2 and influenza co-infections are not well understood [20]. Even if co-infected individuals do not experience increased risks of serious disease, the dual demands on the NHS of caring for COVID-19 and influenza patients is likely to put its resources under pressure. It is therefore essential to monitor ongoing infection risks of both SARS-CoV-2 and influenza so that appropriate steps are in place to protect the health of the public and maintain capacity in the NHS.

In conclusion, during round 15a, we observed the highest prevalence of swab-positivity in England that we have seen to date in REACT-1, with the highest rates being found among school-aged children. Despite the observed fall in the most recent data, these high rates highlight ongoing risks to clinically extremely vulnerable children as well as increasing the risk of infection among close contacts including household members, particularly if they have not been vaccinated [16]. The accelerated vaccination of children aged 12 years and over should help to limit transmission as should ongoing immunisation of previously unvaccinated or single-vaccinated adults, along with the delivery of third doses to vulnerable groups, health and care home workers and older adults.

## Data Availability

Access to REACT-1 data is restricted due to ethical and security considerations. Summary statistics and descriptive tables from the current REACT-1 study are available in the Supplementary Information and at the following link https://github.com/mrc-ide/reactidd/tree/master/inst/extdata/react1_r15_interim. Additional summary statistics and results from the REACT-1 programme are also available at https://www.imperial.ac.uk/medicine/research-and-impact/groups/react-study/real-time-assessment-of-community-transmission-findings/and https://github.com/mrc-ide/reactidd/tree/master/inst/extdata REACT-1 Study Materials are available for each round at https://www.imperial.ac.uk/medicine/research-and-impact/groups/react-study/react-1-study-materials/

## Ethics

We obtained research ethics approval from the South Central-Berkshire B Research Ethics Committee (IRAS ID: 283787).

## Contributors

PE and CAD are corresponding authors. PE, MC-H and CAD conceived the study and the analytical plan. MC-H, OE, BB, HWang, DH and CEW performed the statistical analyses. HWang, OE, DH, BB, and MW curated the data. CA, PJD, DA, WB, GT, GC, HW, AD provided study oversight and results interpretation. AJP generated the sequencing data. AD and PE obtained funding. All authors revised the manuscript for important intellectual content and approved the submission of the manuscript. PE had full access to the data and takes responsibility for the integrity of the data and the accuracy of the data analysis, and for the decision to submit for publication.

## Acknowledgements

MC-H and MW acknowledge support from the H2020-EXPANSE project (Horizon 2020 grant No 874627). MC-H and BB acknowledge support from Cancer Research UK, Population Research Committee Project grant ‘Mechanomics’ (grant No 22184 to MC-H). CAD acknowledges support from the MRC Centre for Global Infectious Disease Analysis and National Institute for Health Research (NIHR) Health Protection Research Unit (HPRU). GC is supported by an NIHR Professorship. HW acknowledges support from an NIHR Senior Investigator Award and the Wellcome Trust (205456/Z/16/Z). PE is Director of the Medical Research Council (MRC) Centre for Environment and Health (MR/L01341X/1, MR/S019669/1). PE acknowledges support from Health Data Research UK (HDR UK); the NIHR Imperial Biomedical Research Centre; NIHR Health Protection Research Units in Chemical and Radiation Threats and Hazards, and Environmental Exposures and Health; the British Heart Foundation Centre for Research Excellence at Imperial College London (RE/18/4/34215); and the UK Dementia Research Institute at Imperial College London (MC_PC_17114). We thank The Huo Family Foundation for their support of our work on COVID-19.

We thank key collaborators on this work – Ipsos MORI: Kelly Beaver, Sam Clemens, Gary Welch, Nicholas Gilby, Kelly Ward, Galini Pantelidou and Kevin Pickering; Institute of Global Health Innovation at Imperial College London: Gianluca Fontana, Justine Alford; School of Public Health, Imperial College London: Eric Johnson, Rob Elliott, Graham Blakoe; Quadram Institute, Norwich, UK: Alexander J. Trotter; North West London Pathology and Public Health England (now UKHSA) for help in calibration of the laboratory analyses; Patient Experience Research Centre at Imperial College London and the REACT Public Advisory Panel; NHS Digital for access to the NHS register; the Department of Health and Social Care for logistic support.

## Supplementary Tables and Figures

**Supplementary Table 1.** Unweighted and weighted prevalence of swab-positivity from REACT-1 across rounds 1 to 15a.

| Round | Tested swabs | Positive swabs | Unweighted prevalence (95% CI) | Weighted prevalence (95% CI) | First sample | Last sample |
| --- | --- | --- | --- | --- | --- | --- |
| 1 | 120,620 | 159 | 0.13% (0.11%, 0.15%) | 0.16% (0.13%, 0.19%) | 01/05/20 | 01/06/20 |
| 2 | 159,199 | 123 | 0.08% (0.07%, 0.09%) | 0.09% (0.07%, 0.11%) | 19/06/20 | 07/07/20 |
| 3 | 162,821 | 54 | 0.03% (0.03%, 0.04%) | 0.04% (0.03%, 0.05%) | 24/07/20 | 11/08/20 |
| 4 | 154,325 | 137 | 0.09% (0.08%, 0.11%) | 0.13% (0.01%, 0.15%) | 20/08/20 | 08/09/20 |
| 5 | 174,949 | 824 | 0.47% (0.44%, 0.50%) | 0.60% (0.55%, 0.71%) | 18/09/20 | 05/10/20 |
| 6 | 160,175 | 1,732 | 1.08% (1.03%, 1.13%) | 1.30% (1.21%, 1.39%) | 16/10/20 | 02/11/20 |
| 7 | 168,181 | 1,299 | 0.77% (0.73%, 0.82%) | 0.94% (0.87%, 1.01%) | 13/11/20 | 03/12/20 |
| 8 | 167,642 | 2,282 | 1.36% (1.31%, 1.42%) | 1.57% (1.49%, 1.66%) | 06/01/21 | 22/01/21 |
| 9 | 165,456 | 689 | 0.42% (0.39%, 0.45%) | 0.49% (0.44%, 0.55%) | 04/02/21 | 23/02/21 |
| 10 | 140,844 | 227 | 0.16% (0.14%, 0.18%) | 0.20% (0.17%, 0.23%) | 11/03/21 | 30/03/21 |
| 11 | 127,408 | 115 | 0.09% (0.07%, 0.11%) | 0.10% (0.08%, 0.13%) | 15/04/21 | 03/05/21 |
| 12* | 108,911 | 135 | 0.12% (0.10%, 0.15%) | 0.15% (0.12%, 0.18%) | 20/05/21 | 07/06/21 |
| 13 | 98,233 | 527 | 0.54% (0.49%, 0.58%) | 0.63% (0.57%, 0.69%) | 24/06/21 | 12/07/21 |
| 14** | 100,527 | 764 | 0.76% (0.71%, 0.82%) | 0.83% (0.76%, 0.89%) | 09/09/21 | 27/09/21 |
| 15a*** | 67,208 | 1,021 | 1.52% (1.43%, 1.61%) | 1.72% (1.61%, 1.84%) | 19/10/21 | 29/10/21 |

\* Sampling strategy changed for round 12 and subsequent rounds. Therefore unweighted prevalence is not directly comparable with previous rounds
\*\* Sample handling changed in round 14. Therefore prevalence is not directly comparable with previous rounds
\*\*\* Sample shipment in round 15 was only through post.

**Supplementary Table 2.** Proportion of each Delta sub-lineage detected in 126 positive samples from round 15a.

| Sub-lineage | N (126) | Proportion |
| --- | --- | --- |
| B.1.617.2 (Delta/Sub-lineage not known) | 17 | 0.135 ( 0.086 , 0.205 ) |
| AY.4 | 76 | 0.603 ( 0.516 , 0.684 ) |
| AY.5 | 4 | 0.032 ( 0.012 , 0.079 ) |
| AY.6 | 1 | 0.008 ( 0.001 , 0.044 ) |
| AY.25 | 1 | 0.008 ( 0.001 , 0.044 ) |
| AY.34 | 1 | 0.008 ( 0.001 , 0.044 ) |
| AY.39 | 2 | 0.016 ( 0.004 , 0.056 ) |
| AY.4.2 | 13 | 0.103 ( 0.061 , 0.169 ) |
| AY.43 | 9 | 0.071 ( 0.038 , 0.130 ) |
| AY.44 | 2 | 0.016 ( 0.004 , 0.056 ) |

\* Note lineage data are only available up to 23 October

**Supplementary Figure 1.**
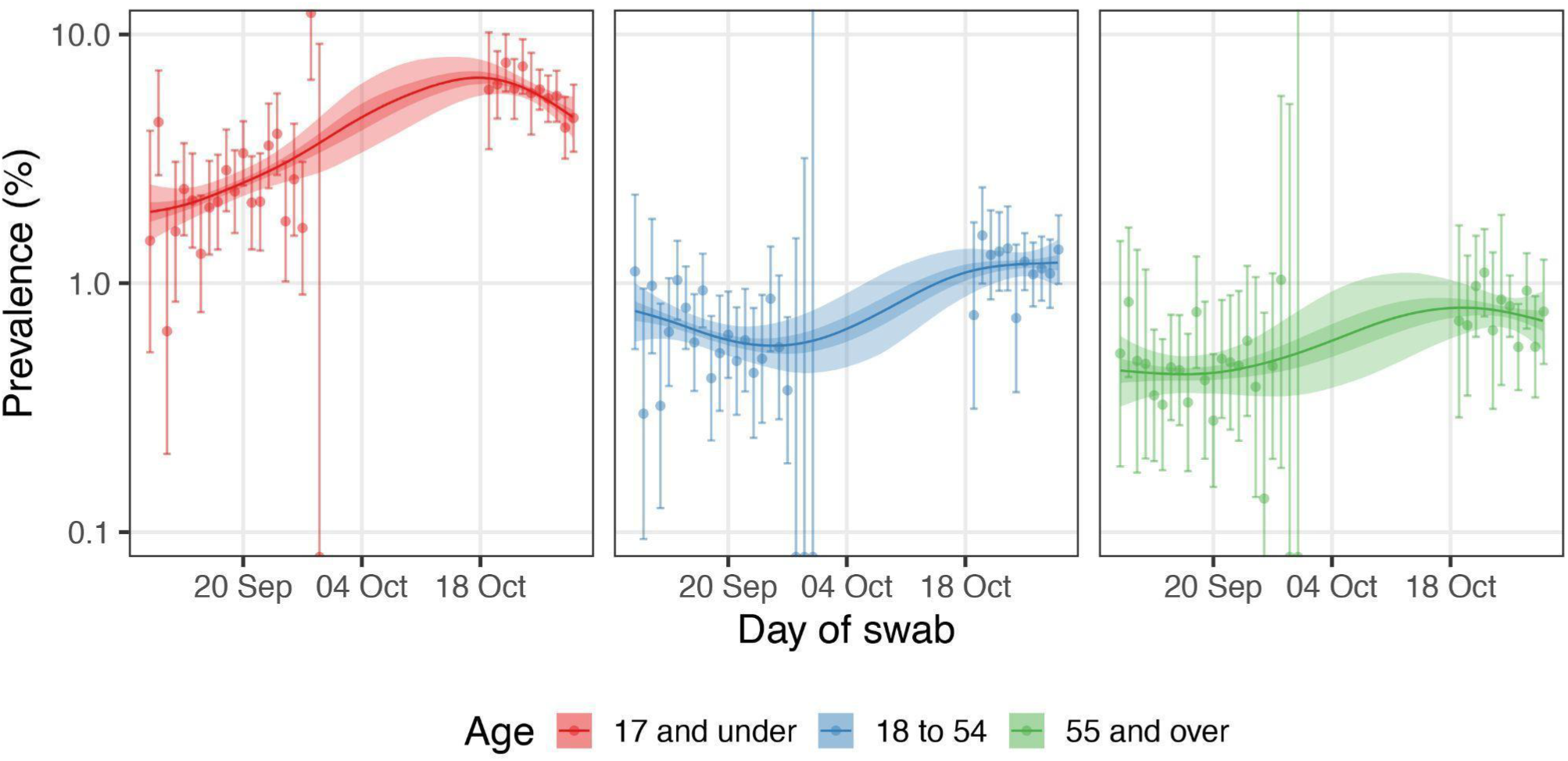
Comparison of P-spline models fit to all rounds of REACT-1 for those aged 17 years and under (red), those aged 18 to 54 years inclusive (blue) and those aged 55 years and over (green). Shown here only for the period of round 14 and round 15a (up to 29 October, 2021). Shaded regions show 50% (dark shade) and 95% (light shade) posterior credible interval for the P-spline models. Results are presented for each day (X axis) of sampling for round 14 and round 15a and the prevalence of swab-positivity is shown (Y axis) on a log scale. Weighted observations (dots) and 95% confidence intervals (vertical lines) are also shown.

## References

1. Riley S, Ainslie KEC, Eales O, Walters CE, Wang H, Atchison C, et al. Resurgence of SARS-CoV-2: Detection by community viral surveillance. Science. 2021;372: 990–995.

2. Riley S, Atchison C, Ashby D, Donnelly CA, Barclay W, Cooke GS, et al. REal-time Assessment of Community Transmission (REACT) of SARS-CoV-2 virus: Study protocol. Wellcome Open Res. 2020;5: 200.

3. Elliott P, Haw D, Wang H, Eales O, Walters CE, Ainslie KEC, et al. Exponential growth, high prevalence of SARS-CoV-2, and vaccine effectiveness associated with the Delta variant. Science. 2021; eabl9551.

4. Chadeau-Hyam M, Wang H, Eales O, Haw D, Bodinier B, Whitaker M, et al. REACT-1 study round 14: High and increasing prevalence of SARS-CoV-2 infection among school-aged children during September 2021 and vaccine effectiveness against infection in England. bioRxiv. 2021. doi:10.1101/2021.10.14.21264965

5. Real-time Assessment of Community Transmission (REACT) study. [cited 3 Nov 2021]. Available: https://www.imperial.ac.uk/medicine/research-and-impact/groups/react-study/

6. Quick J. nCoV-2019 sequencing protocol v3 (LoCost). protocols.io; 25 Aug 2020 [cited 3 Nov 2021]. Available: https://www.protocols.io/view/ncov-2019-sequencing-protocol-v3-locost-bh42j8ye

7. Baker DJ, Aydin A, Le-Viet T, Kay GL, Rudder S, de Oliveira Martins L, et al. CoronaHiT: high-throughput sequencing of SARS-CoV-2 genomes. Genome Med. 2021;13: 21.

8. ncov2019-artic-nf: A Nextflow pipeline for running the ARTIC network’s fieldbioinformatics tools (https://github.com/artic-network/fieldbioinformatics), with a focus on ncov2019. Github; Available: https://github.com/connor-lab/ncov2019-artic-nf

9. pangolin: Software package for assigning SARS-CoV-2 genome sequences to global lineages. Github; Available: https://github.com/cov-lineages/pangolin

10. Team RC. R Core Team R: A language and environment for statistical computing R foundation for statistical computing. Austria, Vienna. 2018.

11. Sharot T. Weighting survey results. 1986 [cited 3 Nov 2021]. Available: http://www.redresearch.com/wp/wp-content/uploads/2016/01/Weighting-Survey-Results.pdf

12. Hoffman MD, Gelman A. The No-U-Turn Sampler: Adaptively Setting Path Lengths in Hamiltonian Monte Carlo. arXiv [stat.CO]. 2011. Available: http://arxiv.org/abs/1111.4246

13. Bi Q, Wu Y, Mei S, Ye C, Zou X, Zhang Z, et al. Epidemiology and transmission of COVID-19 in 391 cases and 1286 of their close contacts in Shenzhen, China: a retrospective cohort study. Lancet Infect Dis. 2020;20: 911–919.

14. Lang S, Brezger A. Bayesian P-Splines. J Comput Graph Stat. 2004;13: 183–212.

15. Riley S, Ainslie KEC, Eales O, Walters CE, Wang H, Atchison C, et al. REACT-1 round 6 updated report: high prevalence of SARS-CoV-2 swab positivity with reduced rate of growth in England at the start of November 2020. bioRxiv. medRxiv; 2020. doi:10.1101/2020.11.18.20233932

16. Singanayagam A, Hakki S, Dunning J, Madon KJ, Crone MA, Koycheva A, et al. Community transmission and viral load kinetics of the SARS-CoV-2 delta (B.1.617.2) variant in vaccinated and unvaccinated individuals in the UK: a prospective, longitudinal, cohort study. Lancet Infect Dis. 2021;0. doi:10.1016/S1473-3099(21)00648-4

17. UK Health Security Agency. Testing at private lab suspended following NHS Test and Trace investigation. In: GOV.UK [Internet]. 15 Oct 2021 [cited 3 Nov 2021]. Available: https://www.gov.uk/government/news/testing-at-private-lab-suspended-following-nhs-test-and-trace-investigation

18. UKHSA. SARS-CoV-2 variants of concern and variants under investigation in England: Technical briefing 27. UKHSA; 2021 Oct. Report No.: 27. Available: https://assets.publishing.service.gov.uk/government/uploads/system/uploads/attachment_data/file/1029715/technical-briefing-27.pdf

19. Weekly national Influenza and COVID-19 surveillance report Week 43 report (up to week 42 data). UKHSA; 2021 Oct. Available: https://assets.publishing.service.gov.uk/government/uploads/system/uploads/attachment_data/file/1029418/Weekly_Flu_and_COVID-19_report_w43.pdf

20. Dadashi M, Khaleghnejad S, Abedi Elkhichi P, Goudarzi M, Goudarzi H, Taghavi A, et al. COVID-19 and Influenza Co-infection: A Systematic Review and Meta-Analysis. Front Med. 2021;8: 681469.

